# High performance of CD4 rapid testing by lay providers in Malawi: Results from a prospective diagnostic accuracy study supporting decentralized advanced HIV disease screening

**DOI:** 10.64898/2026.01.07.26343324

**Authors:** Agness Thawani, Ethel Rambiki, Jacqueline Huwa, Layout Gabriel, Aubrey Kudzala, Christine Kiruthu-Kamamia, Pachawo Bisani, Shameem Buleya, Nathan P Ford, Cheryl C Johnson, Ajay Rangaraj, Robert Luo, Celine Lastrucci, Busisiwe Msimanga, Rose Nyirenda, Bilaal Wilson, Andreas Jahn, Claudia Wallrauch, Rachael Burke, Tom Heller

## Abstract

CD4 testing is essential for identifying people with Advanced HIV Disease to enable provision of a diagnostic package (serum cryptococcal antigen testing and urine lipoarabinomannan testing) and prophylaxis. Lay providers (HIV diagnostic assistants) might be able to perform CD4 testing and advanced HIV diagnostics using lateral flow assays (LFAs). We conducted a prospective diagnostic accuracy study comparing LFA(Visitect) CD4 results performed by HIV diagnostic assistants and by laboratory technicians, using paired quantitative CD4 results from the PIMA device (performed by a nurse) as the reference standard. We also compared results of serum cryptococcal antigen (CrAg) and urine lipoarabinomannan (LAM) tests performed by HDAs to those performed by nurses. Implementation costs were estimated. We recruited 308 participants. Median CD4 was 248 cells/mm^3^; 115 (37.3%) patients had values below 200 cells/mm^3^. Sensitivity and specificity for determining CD4 below 200 cells/mm^3^ using the LFA operated by HIV diagnostic assistants was 94.8% (95%CI 89.1 – 97.6%) and 92.2% (95%CI 87.6 – 95.2%), respectively. Test performance of the LFA performed by laboratory technicians on batch after storage for up to eight hours was substantially worse. Subsequent serum-CrAg and urine-LAM test performed by HIV diagnostic assistants and nurses showed an agreement of 98.1% (kappa=0.74) and 98.1% (k=0.85), respectively. Incremental cost for CD4 test performed by on near patient device based quantitative test was $8.69 and for semi-quantitative LFA by HIV diagnostic assistants was $5.24 (in 2024 US dollars). Trained lay providers can accurately perform CD4, TB-LAM, and CrAg testing. Delaying CD4 testing by batching LFAs at the end of the day led to highly inaccurate results. Our findings support task sharing for decentralized advanced HIV disease testing.

## Introduction

Despite the remarkable progress with antiretroviral therapy (ART) rollout in sub-Saharan Africa, a large proportion of People Living with HIV (PLHIV) continue to present with Advanced HIV Disease (AHD) [1–5]. In 2017, the World Health Organization (WHO) recommended a package of care for AHD, which included screening, treatment or prophylaxis for major opportunistic infections, rapid initiation of ART, and intensified adherence support for all patients presenting with AHD [6]. AHD remains a leading cause of hospital admissions among People Living with HIV globally [7]. Enhanced infection prophylaxis has been shown to reduce mortality in severely immunosuppressed adults and older children initiating ART [8]. Most reporting countries have fully (75%) or partially (17%) adopted this recommended AHD package of care [9].

CD4 testing plays a key role in the identification of AHD due to the large proportion of patients with advanced immune suppression who present with few or no clinical symptoms. People with AHD are at high risk of rapid disease progression and development of opportunistic infections. A CD4 cell count of ≤200 cells/mm3 should trigger testing for extra-pulmonary and disseminated TB using urinary lipoarabinomannan antigen (urine-LAM) and cryptococcal antigen in serum (serum-CrAg) to screen for Cryptococcal meningitis (CM) using lateral flow assays (LFA) [6].

Despite policy recommendations, there is suboptimal uptake of AHD services primarily due to limited access to CD4 testing at, or near, the point-of-care (POC). Access to CD4 testing represents the main bottleneck in the AHD cascade, hindering early identification, further investigation and treatment.

We aimed to evaluate the performance of the CD4 LFA test conducted by HIV diagnostic assistants (HDAs). The primary objective was to measure the accuracy and feasibility of using CD4 LFA when performed and interpreted by HDAs, compared with the current standards performed by trained and registered nurses and laboratory technicians. Secondly, we assessed if HDAs can correctly perform and interpret serum-CrAg and urine-LAM tests for CM and disseminated TB respectively, and to compare the test performance with existing published data. Finally, we report estimated potential cost savings due to task sharing for our setting.

## Methods

### Study setting and population

Two HIV diagnostic assistants (HDAs) were recruited for this study. They were trained and certified in HIV testing, but without prior experience with the CD4 LFA, urine LAM, or serum CrAg testing. The HDA is a novel HIV testing cadre in Malawi, with early findings showing feasibility and effectiveness in delivering HIV testing services [10]. The HDAs received a half-day training in advanced HIV disease, TB, and cryptococcal meningitis followed by a 5-day practical training in the laboratory. HDAs were also oriented to conduct tests using the WHO-prequalified VISITECT® CD4 rapid assay, consistent with international standards [11].

This Cross-section study was conducted at the Lighthouse ART Clinic at Kamuzu Central Hospital in Lilongwe, Malawi, between 18^th^ August 2023 and 08^th^ March 2024. Lighthouse ART clinic is a large and well-established HIV clinic based at Kamuzu Central Hospital in Lilongwe, Malawi, and has played a central role in the country’s HIV response for over 15 years [12]. The clinic runs a dedicated mini laboratory for treatment monitoring and diagnosis of advanced HIV disease. It is staffed by laboratory technicians and nurses who perform CD4 testing on venous EDTA samples using a quantitative near point-of-care device-based test. Patients presenting with AHD are managed according to the Malawi National guidelines, which recommend serum CrAg and urine LAM testing for individuals with CD4 ≤200 cells/µL [13].

### Tests used

The semi-quantitative LFA used was *VISITECT®CD4 test* product (Accubio, UK) [11,14), the quantitative near point of care device-based test was the PIMA CD4 test machine (Abbott, USA). Serum CrAg tests were from IMMY (USA), and the urine LAM test was Determine LAM (Abbott, USA). All tests were either WHO prequalified or WHO recommended and were performed according to the manufacturer’s instructions and local standard operating procedures. For urine LAM tests, results were as positive (band as dark or darker than the reference card line) or negative (no band or faint band lighter than the reference card line). Grades were not recorded.

#### Eligibility criteria

All PLHIV aged ≥ 18 years who were either initiated on ART or were referred to the lab for CD4 testing were eligible. Referral reasons for CD4 testing included new HIV diagnosis, high viral load, re-engagement in ART after interruption of >2 months, or diagnosis of a WHO stage 3 or 4 clinical condition. Sequential individuals requiring CD4 testing were approached for written informed consent. Exclusion criteria included age <18 years and being too ill for outpatient care.

#### Study design and procedures

After consenting for study, participants were first seen by an HAD who collected a finger-prick blood sample and immediately performed CD4 LFA testing in a separate consultation room. For patients with CD4 counts ≤200 cells/mm³ on LFA, HDAs also performed serum-CrAg[15] and urine-LAM [16] testing. HDAs collected a second capillary blood sample from a finger-prick for the serum-CrAg tests, and patients provided a self-collected urine specimen for the LAM tests. Test results were interpreted and documented on a standard form by the HDAs. All participants were then referred to the mini laboratory, where a nurse collected a 5 mL venous blood sample in an EDTA tube. The nurse performed CD4 testing using a near point-of-care (POC) device. The nurse was blinded to the LFA CD4 results and independently recorded the device-based results on a separate form. For participants with CD4 counts ≤200 cells/mm³, the nurse also performed serum cryptococcal antigen (CrAg) and urine lipoarabinomannan (LAM) tests.

Finally, the remaining blood in the EDTA tube was stored in the mini-laboratory fridge, and in some cases for up to eight hours, after which a laboratory technician performed CD4 LFA testing on these samples.

All individual results were recorded on a study form, which was put in a sealed envelope and collected by a data clerk. HIV diagnostic assistants, nurses and laboratory technicians were blinded to all results obtained by the others. Only results obtained by nurses were shared with the treating clinician for patient management.

#### Data analysis

We calculated CD4 LFA sensitivity, specificity, negative predictor value (NPV) and positive predictor value (PPV) with an assumption that the near device based near POC CD4 measure was a perfect reference standard. However, we know what PIMA CD4 machines can vary by up to 15% or 30 cells/mm^3^ at CD4 counts under 200 cells/mm^3^. We therefore did two sensitivity analyses. We did a sensitivity analysis under “best case scenario” where all LFA results were assumed to be correct if the near POC device result was within 170 to 230 cells/mm^3^. We did not have enough data points for latent class analysis allowing for reference standard inaccuracy and conditional dependence between reference standard and index text results. We estimated NPV and PPV at advanced HIV disease prevalence of 40% (observed in this study) and at 28% (the estimated AHD prevalence for Malawi).

A minimum sample of 308 patient sample pairs was deemed adequate to determine inter-rater agreement of ≥90% (Kappa) assuming 45% of patients had a CD4≤200 cells/mm^3^ as was previously seen at lighthouse [17].

For the costs, we compared the incremental cost per test of the different test and staffing modalities (device based vs. LFA; diagnostic assistant vs. nurse) in 2023 US dollars. Costs of consumables were based on unit prices, including procurement and shipping charges (Ministry of Health logistics data). Staff cost was estimated using the national wages at entry level of Malawi MOH salary scale and the average duration of the testing procedures. We did not include fixed device costs, quality assurance / quality control costs or training costs.

#### Ethical approval

The study protocol together with informed consent forms and data collection tools were approved by the Malawi National Health Sciences Research Committee (NHSRC Reference Number: 2881). Written informed consent was obtained from all participants. Participants who were illiterate provided a thumb print on the form to indicate their consent to participate.

## Results

Between 18 August 2023 and 08 March 2024, we enrolled 308 participants. The median age was 39 years [IQR 31-48]; 167 (54.5%) were female. Median CD4 count in the samples determined by PIMA^®^ was 248 cells/mm^3^ [IQR 136-483]; 115 of the patients (37.3%) had CD4 cell counts ≤200 cells/mm^3^.

### Performance of CD4 LFA tests compared to near point of care device-based test

CD4 was performed on all 308 participants by both HIV diagnostic assistants (using LFA) and nurses (using near point of care device-based tests). One person did not get a result for LFA from a lab technician (figure 1).

**Figure 1:**
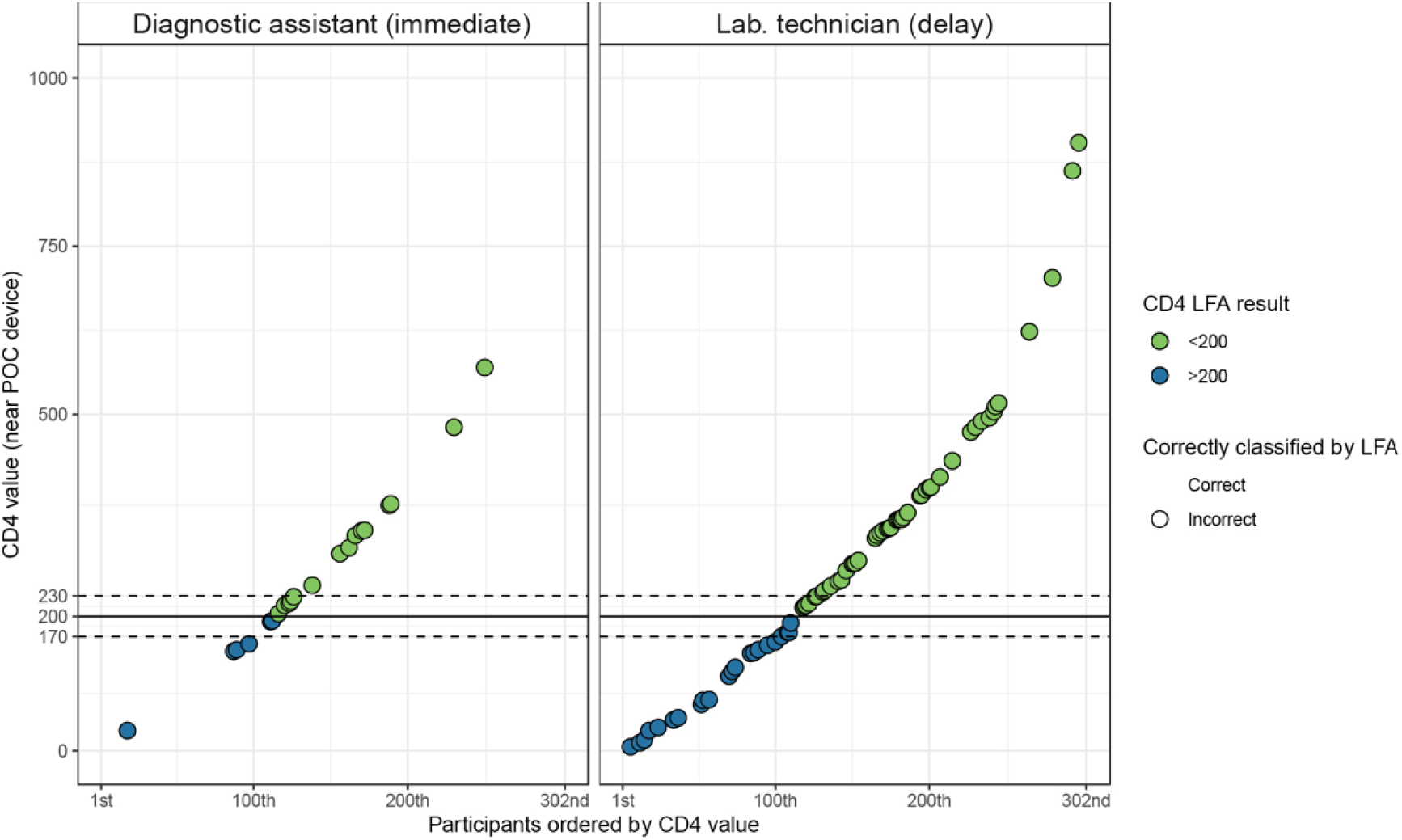
CD4 test results by near point of care device and LFA. **Figure 1 legend**: CD4 counts by near point of care quantitative device performed by nurses (CD4 count on y axis) and by semiquantitative LFA by HIV diagnostic assistants (color of point). Horizontal lines indicate various CD4 cell counts. Light shaded dots show concordant results; discordant results are shown as dark shaded dots. Results for six participants with CD4 >1000 (all concordant) are not shown on graph.

Cross-tabulated outcomes shown in table 1. If the true population prevalence of advanced HIV disease was 28%, the negative predictor value and positive predictor values for LFA by HIV diagnostic assistants would be 0.83 (95% CI: 0.74 – 0.89) and 0.98 (95% CI: 0.95-0.99), respectively. Supplementary Table 1 shows the results of a sensitivity using a ‘best case scenario’ in which all LFA results with near POC device values within 15% of the threshold are considered correct. Test performance was substantially worse for residual EDTA samples tested at the end of the day on batch.

**Table 1:** Diagnostic accuracy LFA CD4 compared to device based near point of care CD4.

| Index test | Near POC device test<br>(reference standard) |  |  |
| --- | --- | --- | --- |
|  | Reference test<br><200<br>cells/mm<sup>3</sup> | Reference test<br>>200<br>cells/mm<sup>3</sup> |  |
| LFA test on fresh capillary blood by HIV diagnostic assistants. |  |  |  |
| Index test <200 cells/mm<sup>3</sup> | 109 | 15 | PPV: 0.88 (0.81-9.93) |
| Index test >200 cells/mm<sup>3</sup> | 6 | 178 | NPV: 0.97 (0.93 – 0.99) |
|  | Sensitivity:<br>0.95 (0.89 – 0.98) | Specificity:<br>0.92 (0.88 – 0.96) |  |
| LFA test on residual EDTA samples after delay by laboratory technicians |  |  |  |
| Index test <200 cells/mm<sup>3</sup> | 93 | 47 | PPV: 0.66 (0.58-0.74) |
| Index test >200 cells/mm<sup>3</sup> | 22 | 145 | NPV: 0.87 (0.81 – 0.92) |
|  | Sensitivity:<br>0.81 (0.72-0.88) | Specificity:<br>0.76 (0.69-0.81) |  |

**Legend:** PPV: Positive predictive value, NPV: negative predictive value (at observed prevalence)

### Results of serum-CrAg and urine-LAM

Of the 115 individuals with LFA CD4 <200 cells/mm³, 107 underwent CrAg testing by both the nurse and the HDA, while 105 underwent LAM testing by both cadres. Two individuals were missing CrAg test results, and three were missing LAM test results from the nurses. Additionally, five individuals were found to have CD4 counts >200 cells/mm³ based on Visitect testing conducted by the HDAs and were therefore deemed ineligible for CrAg and LAM testing.

Among the 193 participants with CD4 counts >200 cells/mm³ measured using near point-of-care (POC) devices, five had serum CrAg tests and seven had urine LAM tests performed by nurses.

Among the 107 individuals who had serum CrAg testing performed by both a diagnostic assistant and a nurse, 5 tested CrAg positive according to the nurses, of whom 3 were also identified as positive by the HIV diagnostic assistants. There was 100% concordance for negative results. Overall agreement was 98% (Cohen’s kappa = 0.75). In 105 participants where urine LAM was performed by both a HIV diagnostic assistants and nurses, 97 had negative LAM test result by both, Six positive LAM by both and two positive LAM by nurses but negative by HIV diagnostic assistants. Overall agreement was 98% (Cohen’s kappa = 0.85). One person had a positive LAM by nurses and not done by diagnostic assistant, and three people had positive LAM by diagnostic assistant but not done by nurses (table 2).

**Table 2:**
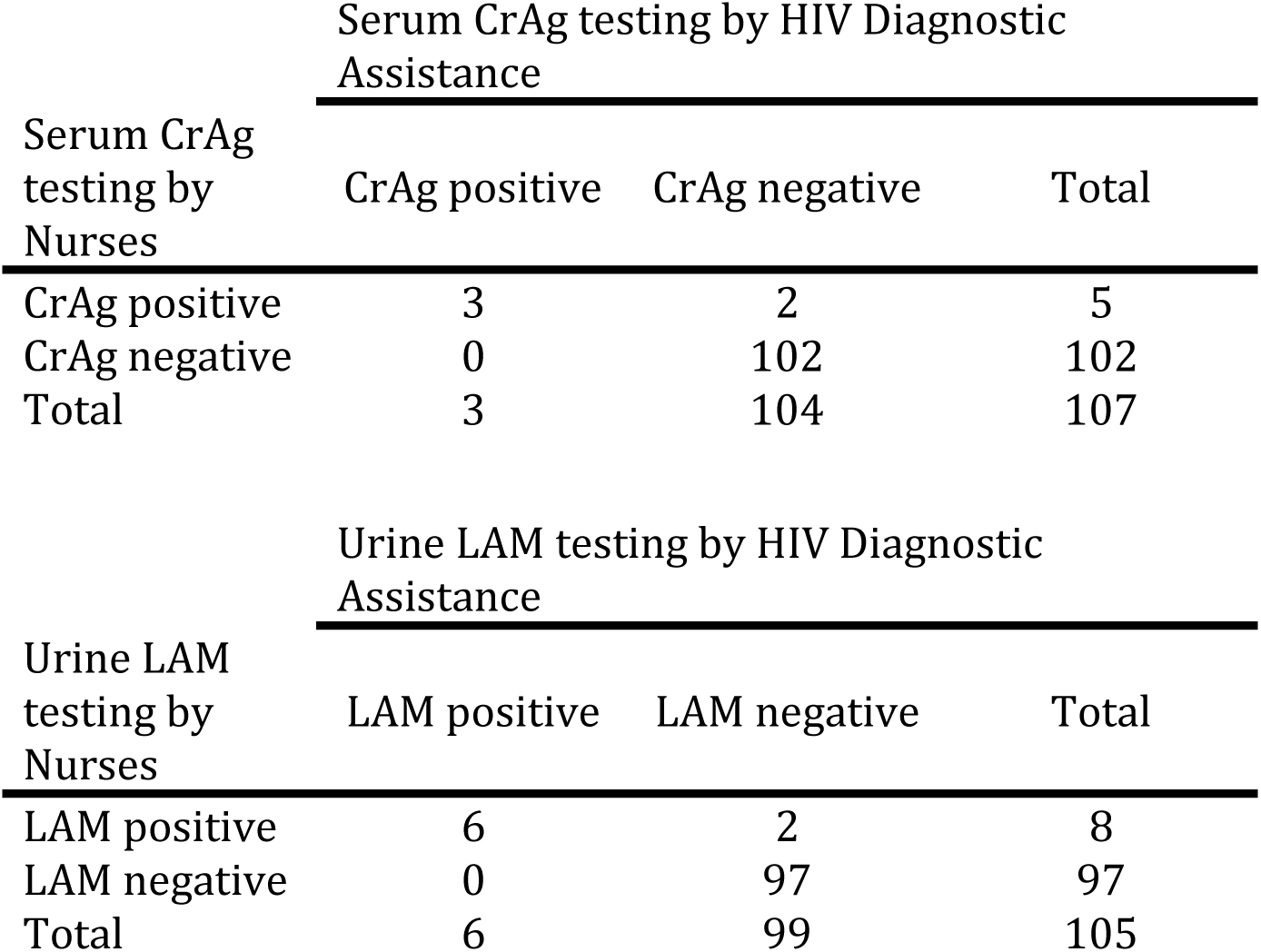
Comparison of urine LAM and serum CrAg testing.

### Cost

We estimated that per-test costs of consumables to Malawi HIV programme (using low-income country preferential pricing) in 2024 US dollars were $7.78 for the device based near POC test and $4.17 for CD4 LFA. The near POC device took 35 minutes of staff time (including time for preparation, blood draw and documentation) and the LFA took 60 minutes. The cost for 35 minutes of nurse staff time was $0.91 and for 60 minutes of diagnostic assistant time was $1.07. Overall incremental cost per CD4 test was $8.69 for near POC device quantitative test by a nurse and $5.24 for semi-quantitative LFA by a diagnostic assistant.

## Discussion

The WHO advanced HIV disease guidelines suggested programs consider task sharing for performing the advanced HIV point-of-care diagnostics although an evidence gap was noted, particularly with more complex assays. Our results show good performance of CD4 LFA in the hands of lay personnel with only a few days of targeted training and it was feasible to task shift to HIV diagnostic assistants. By contrast, results from LFA testing after several hours storage at fridge temperature were substantially less reliable – even when performed by a more experienced cadre of staff (lab technicians).

Overall, the performance characteristics for CD4 by HIV diagnostic assistants on fresh capillary blood we found are compatible with results from previously published reports. Seven studies allowing individual patient data extraction on LFA test performance were identified in the literature [18–24]; the diagnostic performance in our study was at the upper range of sensitivity (62^20^-97^18^%) and specificity (71^24^-95^18^%) of previous evaluations. We attribute the poor performance of LFAs on stored / residual blood to being most likely related to pre-analytic factors such as sample degradation over time rather than poor interpretation by lab technicians. We urge those implementing LFA role out to consider pre-analytic factors.

Overall, the performance characteristics of CD4 testing by HIV diagnostic assistants on fresh capillary blood in our study are consistent with previously published reports. Seven studies providing individual patient data on LFA test performance were identified in the literature [18–24]. The diagnostic performance observed in our study was at the upper range of reported sensitivity (62^20^-97^18^%) and specificity (71^24^-95^18^%) from previous evaluations. We attribute the lower performance of LFAs on stored or residual blood primarily to pre-analytic factors, such as sample degradation over time, rather than errors in interpretation by laboratory technicians. We recommend that programs implementing LFA rollout carefully consider these pre-analytic factors to optimize test accuracy.

We are confident that using HIV diagnostic assistants to perform CD4 and other advanced HIV diagnostic tests is feasible without reducing quality compared to qualified nurses. This confirms the great potential for decentralizing and increasing AHD screening coverage in Malawi and comparable settings. Task-sharing has played a critical role in Malawi’s HIV testing program over the years by enabling far more PLHIV to be diagnosed and started on ART than would have otherwise been possible. The same strategy can now be used to accelerate the scale-up of advanced HIV services.

Whilst we have shown that HIV diagnostic assistants with minimal training were able to use the CD4 LFA test, the current Visitect lateral flow test takes longer than other device-based tests (60 minutes compared to 30 minutes) and has more complicated steps such as adding buffer at specific time points. These time-sensitive steps make the test less suitable for “batch” testing (i.e., one person running multiple tests at the same time) and for completing other tasks while waiting for the next step in the LFA test. There is also limited experience and guidance on quality assurance and quality control for the Visitect LFA, compared to PIMA where QC cartridges and user-friendly instructions for use are readily available. We have shown that under some circumstances (long delays between sample collection and result generation), Visitect results can be very unreliable, highlighting the need for ongoing training and support for testing personnel as part of quality assurance measures.

Patients with advanced HIV disease carry a high burden of morbidity and mortality. Studies have shown that screening tests such as urine-LAM [25] and prophylactic treatments [8] can decrease this mortality, which has led to clear guidelines of advanced HIV disease care [6]. Advanced HIV disease is defined by CD4 cell count, and so determining CD4 levels is the essential entry point for advanced HIV disease care. Our results show excellent performance of the CD4 LFA in the hands of lay testers, with short and targeted training. Serum-CrAg and the urine-LAM results showed a high concordance between the results of nurses and HDAs, but the low number of positive samples limited the precision of the estimates for diagnostic accuracy. Our results are consistent with previously published reports from Lesotho, showing that CrAg LFA screening by lay cadres is feasible^26^. Field implementation evaluations in rural Malawi also suggest that trained HIV diagnostic assistants can perform and interpret rapid tests for opportunistic infections and refer presumed AHD patients to clinicians [27]. There was less concordance in urine LAM readings than serum CrAg readings – presumably because reading a urine LAM test can be subjective when it comes to comparing faint lines with the reference card.

Our cost estimates indicate that using the CD4 LFA test in this setting could result in up to 30% incremental cost savings, with an additional 13% savings achievable by task sharing testing to HDAs. In addition to these cost savings, the LFA facilitates the expansion of advanced HIV diagnostic services to smaller facilities. This is particularly relevant in a decentralized ART program such as Malawi’s [12], where approximately 835 treatment sites provide ART nationwide. In many of these sites, electricity is unreliable or unavailable, healthcare staff are limited, and laboratory technicians are absent. Under these conditions, it is unlikely that nurses, who generally manage these smaller ART clinics, would be able to perform the CD4 LFA test due to competing responsibilities.

HIV diagnostic assistants were introduced in the Malawian HIV program to task-share HIV testing using simple algorithms at the point of care [9–10]. It is therefore sensible to consider using this lay cadre for additional advanced HIV disease (AHD) screening tests. However, because these tests must meet specific quality standards, existing policies on lay provider testing may need to be adapted to strengthen quality control measures. The CD4 LFA test may be less suitable for busy HIV referral clinics with high patient volumes due to the long incubation period and the time-sensitive repeated application of buffer. Delivering a single-day “one stop shop” care to multiple patients in parallel is more challenging with the LFA, which requires approximately 45 hands-on minutes, compared with 20 hands-on minutes for device-based near POC CD4 testing. In clinical settings with better-trained staff and potential inpatient care [28], knowing CD4 counts below 100 cells/mm³ or even below 50 cells/mm³ can inform differential diagnoses and guide therapeutic decisions. In contrast, deploying lay cadres in smaller peripheral facilities with fewer patients, where skilled health workers are occupied with clinical duties, appears to be more feasible. In such settings, patients can be screened and receive same-day treatment for any AHD condition identified. This approach may also be advantageous in sites with unreliable electricity, which limits the use of traditional CD4 machines.

A limitation of our study is that the HIV diagnostic assistants were full-time and focused solely on AHD testing; they did not perform other tests such as HIV, syphilis, or hepatitis B screening as would typically occur in most health facilities. This dedicated focus may have contributed to their excellent performance.

## Conclusion

Lay providers, with brief training, can deliver advanced HIV testing services using LFAs for CD4, urine-LAM, and serum CrAg with accuracy comparable to trained nurses. These findings support the decentralized implementation of advanced HIV disease testing by lay providers, particularly in rural clinics that see few patients per day, lack other CD4 testing devices, or have unreliable electricity and limited laboratory facilities. This implementation model appears highly feasible, potentially cost-saving, and could play a critical role in expanding access to advanced HIV disease testing and treatment, with significant population-level impact.

## Data Availability

All data produced in the present study are available upon reasonable request to the authors

## Acknowledgments

We thank the Ministry of Health, Malawi, for their collaboration and support in implementing this study. We are grateful to all Lighthouse health facility staff, study team and study participants for their contributions.

**Writing – original draft**: AT, ER, TH, RB

**Formal analysis:** AT, ER, AJ, CW, TH, RB

**Conceptualization:** AT, ER, JH, AJ, CW, TH

**Methodology:** AT, ER, JH, AJ, CW, TH

**Investigation:** AT, ER, JH, LG, PB, SB, AJ, CW, TH, RN

**Data curation:** AT, ER, JH, AK, PB, TH

**Validation:** JH, CKK, PB

**Supervision:** AT, ER, JH, LG, CKK, TH

**Project administration:** AT, ER, JH, TH

**Resources:** SB, NF, CJ, AR, RL, CL, BM, RN

**Funding acquisition:** NF, CJ, AR, RL, CL, BM, RN

**Writing – review & editing:** All authors

## Supporting Information Captions

**Supplementary Table 1.**
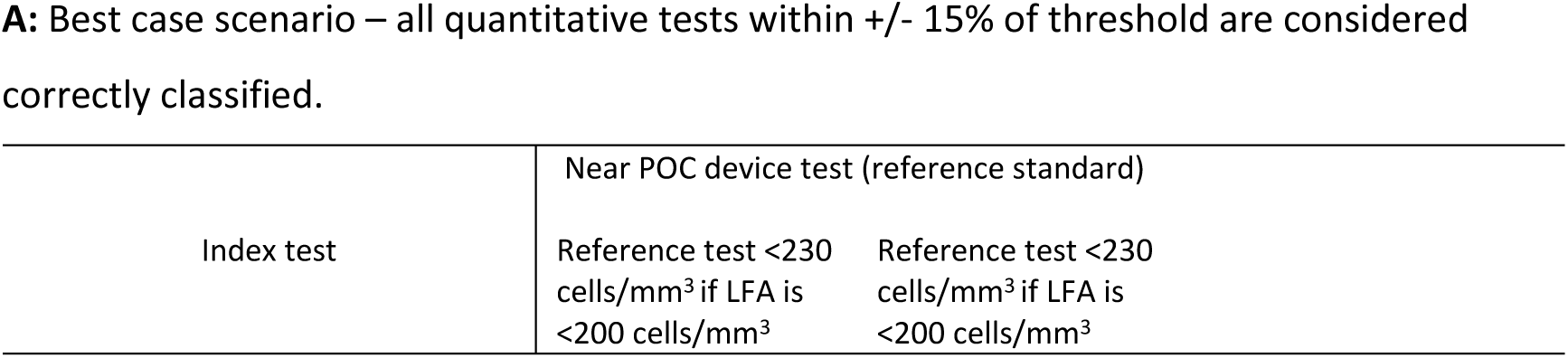
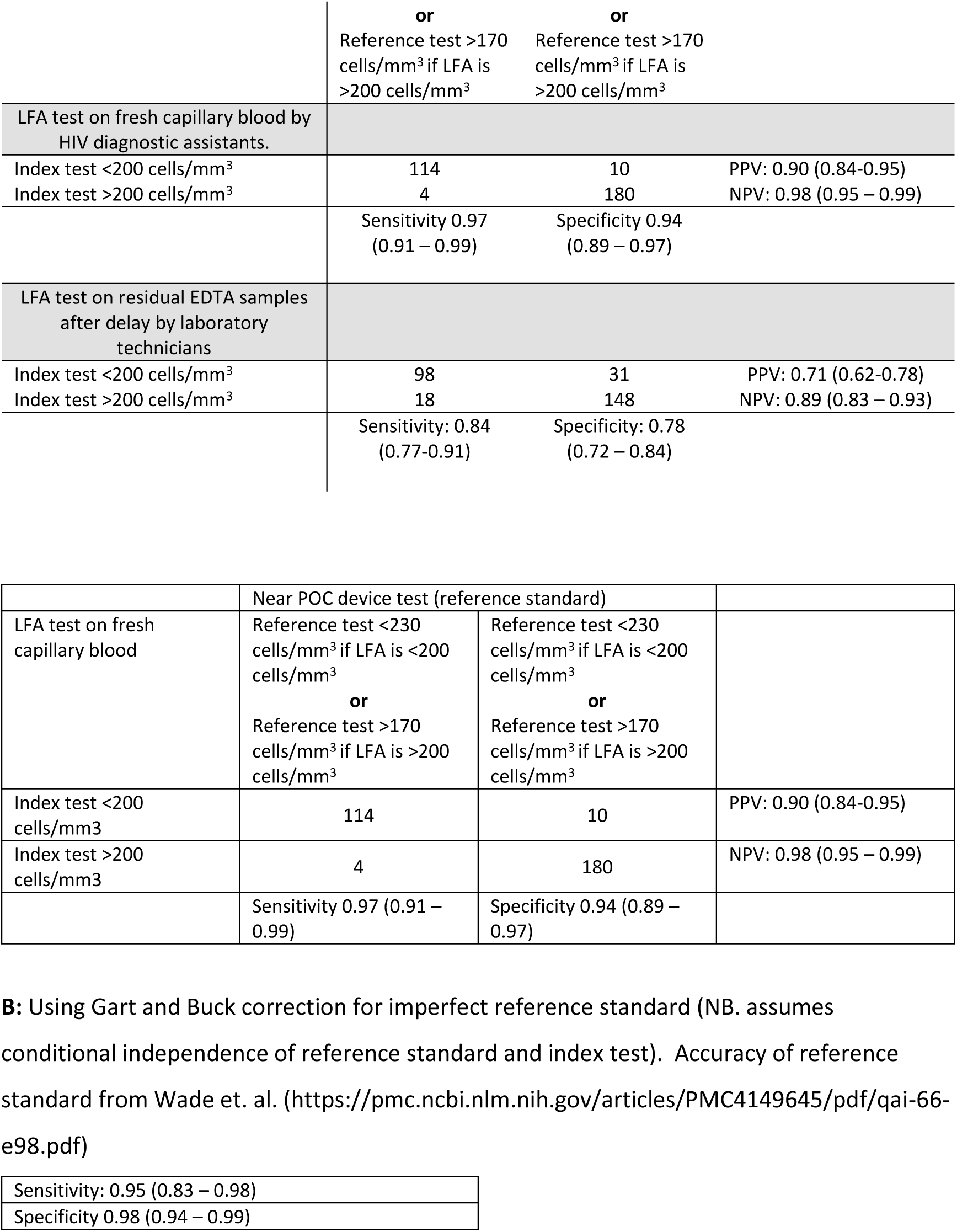
Sensitivity estimates for CD4 testing. Supplementary table 1: Sensitivity estimates for CD4 testing

| Index test | Near POC device test (reference standard) |  |
| --- | --- | --- |
|  | Reference test <230 cells/mm<sup>3</sup> if LFA is <200 cells/mm<sup>3</sup> | Reference test <230 cells/mm<sup>3</sup> if LFA is <200 cells/mm<sup>3</sup> |

|  | or<br>Reference test >170<br>cells/mm<sup>3</sup> if LFA is<br>>200 cells/mm<sup>3</sup> | or<br>Reference test >170<br>cells/mm<sup>3</sup> if LFA is<br>>200 cells/mm<sup>3</sup> |  |
| --- | --- | --- | --- |
| LFA test on fresh capillary blood by<br>HIV diagnostic assistants. |  |  |  |
| Index test <200 cells/mm<sup>3</sup> | 114 | 10 | PPV: 0.90 (0.84-0.95) |
| Index test >200 cells/mm<sup>3</sup> | 4 | 180 | NPV: 0.98 (0.95 – 0.99) |
|  | Sensitivity 0.97<br>(0.91 – 0.99) | Specificity 0.94<br>(0.89 – 0.97) |  |
| LFA test on residual EDTA samples<br>after delay by laboratory<br>technicians |  |  |  |
| Index test <200 cells/mm<sup>3</sup> | 98 | 31 | PPV: 0.71 (0.62-0.78) |
| Index test >200 cells/mm<sup>3</sup> | 18 | 148 | NPV: 0.89 (0.83 – 0.93) |
|  | Sensitivity: 0.84<br>(0.77-0.91) | Specificity: 0.78<br>(0.72 – 0.84) |  |

|  | Near POC device test (reference standard) |  |  |
| --- | --- | --- | --- |
| LFA test on fresh<br>capillary blood | Reference test <230<br>cells/mm<sup>3</sup> if LFA is <200<br>cells/mm<sup>3</sup><br>or<br>Reference test >170<br>cells/mm<sup>3</sup> if LFA is >200<br>cells/mm<sup>3</sup> | Reference test <230<br>cells/mm<sup>3</sup> if LFA is <200<br>cells/mm<sup>3</sup><br>or<br>Reference test >170<br>cells/mm<sup>3</sup> if LFA is >200<br>cells/mm<sup>3</sup> |  |
| Index test <200<br>cells/mm<sup>3</sup> | 114 | 10 | PPV: 0.90 (0.84-0.95) |
| Index test >200<br>cells/mm<sup>3</sup> | 4 | 180 | NPV: 0.98 (0.95 – 0.99) |
|  | Sensitivity 0.97 (0.91 –<br>0.99) | Specificity 0.94 (0.89 –<br>0.97) |  |

|  |
| --- |
| Sensitivity: 0.95 (0.83 – 0.98) |
| Specificity 0.98 (0.94 – 0.99) |

**A:** Best-case scenario, in which all quantitative tests within ±15% of the threshold are considered correctly classified. Sensitivity, specificity, positive predictive value (PPV), and negative predictive value (NPV) are shown for: (i) LFA testing on fresh capillary blood by HIV diagnostic assistants and (ii) LFA testing on residual EDTA samples after delay by laboratory technicians. Reference standard thresholds: <230 cells/mm³ if LFA <200 cells/mm³, and >170 cells/mm³ if LFA >200 cells/mm³.

**B:** Sensitivity and specificity estimates using the Gart and Buck correction for imperfect reference standards, assuming conditional independence between the reference standard and index test. Accuracy of the reference standard based on Wade et al. (2015).

**Supplementary figure 1:**
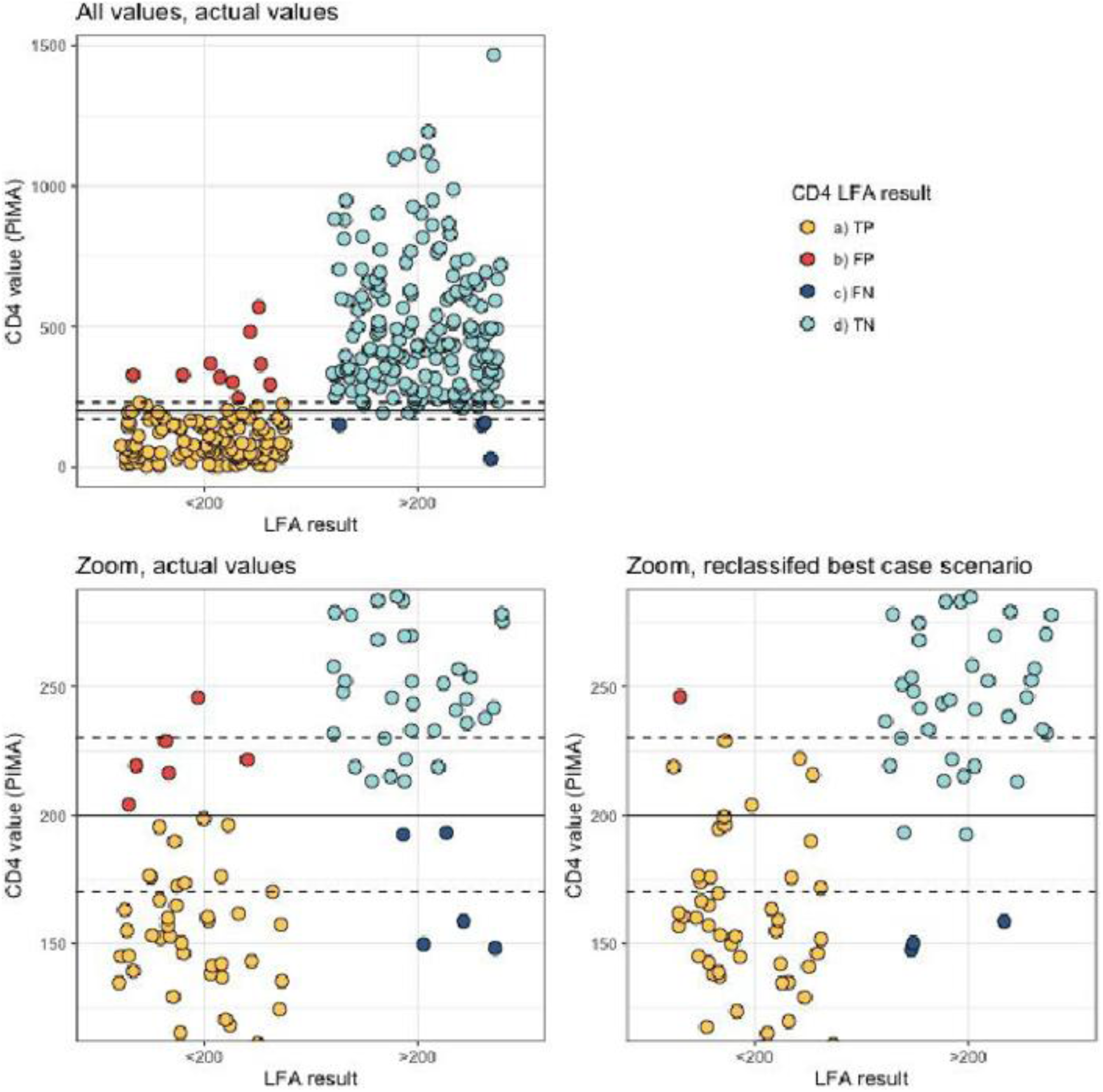
Reclassification of points for “best case scenario”. **Legend:** Top panel shows all results and classification (color). Bottom left hand panel shows results in range 120 to 180 CD4 cells/mm^3^ and their classification using actual values (i.e. assuming no measurement error in reference standard). All tests where LFA is <200 cells/mm^3^ and near POC device if above 200 cells/mm^3^ are considered false positive (orange colored). Bottom right hand panel shows classification using “best case scenario”, where four results where LFA was <200 cells/mm^3^ and near POC device was above 200 cells/mm^3^ but below 230 cells/mm^3^ are reclassified as true positive (yellow colored). Similarly, two results where LFA showed >200 cells/mm^3^ but near POC device was between 170 cells/mm^3^ and 200 cells/mm^3^ are reclassified as true negative (colored light blue).

**Supplementary Figure 2:**
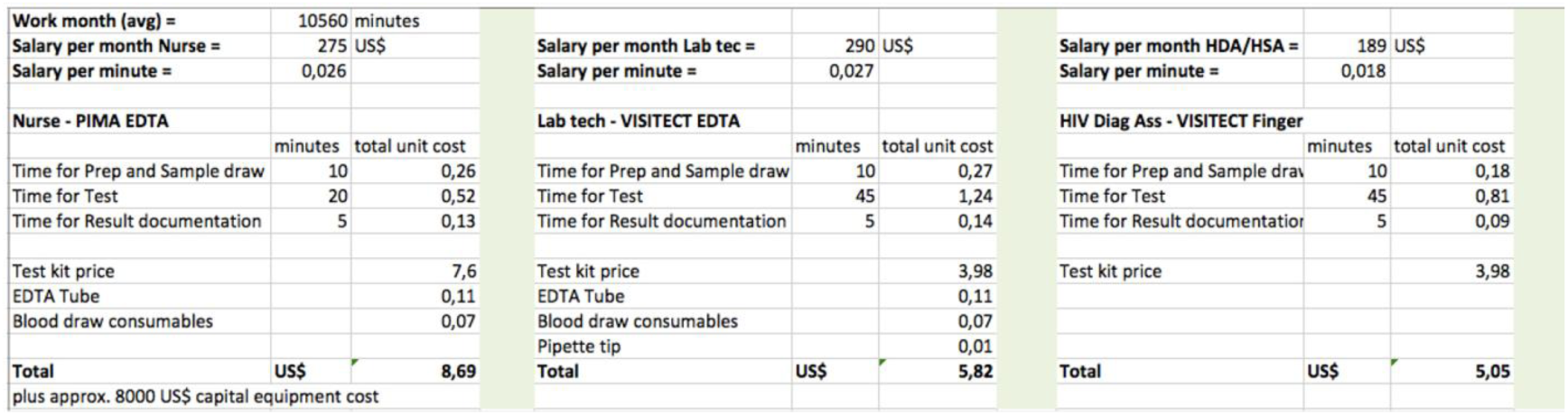
Estimate of costs per CD4 result assuming Malawian 2024 consumable costs and Ministry of Health entry-level wage estimates for various cadres.

### Supplement 3: Literature search for previous published VISITECT test results

*Search criteria:* Literature was searched in PubMed and Google^®^ Scholar using the search string “CD4” AND “point-of-care” AND (“lateral flow assay” OR “VISITECT”) to identify studies assessing test characteristics of the Visitect test. Subsequently a targeted Google^®^ search using the same terms was performed to find relevant conference poster presentations not registered in the abovementioned databases. All studies, which allowed extraction of individual patient data from the published report or poster were included, summarized and analyzed using a random-effects model. Studies using the VISITECT^®^ CD4 350 test were included as it functions on the same test principle.

*Comparison of study results with previously reported test performance:* Seven studies allowing individual patient data extraction on Visitect test performance were identified in the literature^18-24^. Two studies^20,24^ reported disaggregated results for different sample types (EDTA vs. whole blood from finger prick); data was segregated accordingly. All published studies were performed in the lab setting except Obutu 2022^23^ and Kirungi 2022^18^, where VISITECT tests were performed by not further specified facility health care workers and Luchers 2019 (VISITECT finger-prick tests were performed by nurses). The results are summarized in Supplementary Figure 2. Sensitivity ranged from 62^20^-97^18^%, specificity from 71^24^-95^18^%.

Attempting a summary despite the high heterogeneity, overall sensitivity from all studies including our own data was 89% [95%CI 84-93%]; overall specificity 85% [95%CI 81-89%].

**Supplementary Figure 3:**
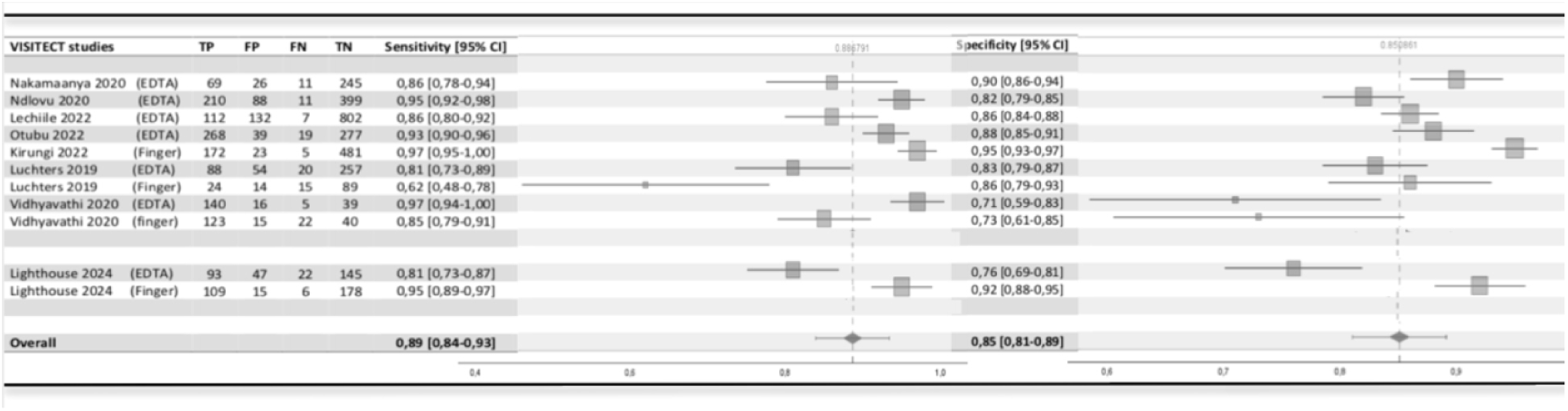
Forest plot of published sensitivity and specificity data of VISITECT^®^ CD4 tests in comparison to results from Lighthouse study. Blood specimen types are specified in the table. Heterogeneity: Sensitivity: χ^2^=54, df=10, p<0.001, *I*^2^=92%; Specificity χ^2^=79, df=10, p<0.001, *I*^2^=91%

## Notes

### Competing Interest Statement

The authors have declared no competing interest.

### Funding Statement

This study was funded by World Health Organisation (WHO)

### Author Declarations

The study protocol together with the informed consent forms was approved by the Malawi National Health Sciences Research Committee which is registered with the USA office for Human Research Protection (OHRP) as an international IRB. IRB number: IRB 00003905 FWA 00005976

## References

1. Carmona S, Bor J, Nattey C, et al. Persistent high burden of advanced HIV disease among patients seeking care in South Africa’s National HIV program. Clin Infect Dis 2018; 66 (suppl2): S111–17.

2. Benzekri NA, Sambou JF, Ndong S, et al. Prevalence, predictors, and management of advanced HIV disease among individuals initiating ART in Senegal, West. BMC Infect Dis 2019; 19:261.

3. Lifson AR, Workneh S, Hailemichael A, et al. Advanced HIV disease among males and females initiating HIV care in rural Ethiopia. J int Assoc Provid AIDS Care 2019; 18:1–7

4. Blankley S, Gashu T, Ahmad B, Belaye AK, Ringtho L, Mesic A, Zizhou S, Casas EC. Lessons learned: Retrospective assessment of outcomes and management of patients with advanced HIV disease in a semi-urban polyclinic in Epworth, Zimbabwe. PLoS One. 2019 Apr 10;14(4):e0214739. doi: 10.1371/journal.pone.0214739. PMID: 30969987; PMCID: PMC6457534.

5. Van der Kop ML, Thabane L, Awiti PO, et al. Advanced HIV disease at presentation to care in Nairobi, Kenya: Late diagnosis or delayed linkage to care? – a cross sectional study. BMC Infect Dis 2016; 16:169.

6. WHO. Guidelines for managing advanced HIV disease and rapid initiation of antiretroviral therapy. July 2017.

7. Ford, Nathan, et al. “Causes of hospital admission among people living with HIV worldwide: a systematic review and meta-analysis.” The lancet HIV 2.10 (2015): e438–e444.

8. Hakim, J., et al. “Enhanced infection prophylaxis reduces mortality in severely immunosuppressed HIV-infected adults and older children initiating antiretroviral therapy in Kenya, Malawi, Uganda and Zimbabwe: the REALITY trial.” (2016).

9. https://cdn.who.int/media/docs/default-source/hq-hiv-hepatitis-and-stis-library/who-hiv-policy-adoption-in-countries_2023.pdf?sfvrsn=e2720212_1

10. Flick, Robert J., et al. “The HIV diagnostic assistant: early findings from a novel HIV testing cadre in Malawi.” AIDS 33.7 (2019): 1215–1224.

11. https://extranet.who.int/prequal/sites/default/files/whopr_files/PQDx_0384-077-00_VISTECT-CD4_AdvancedDisease_v5.0.pdf, accessed 12.4.24

12. Phiri, S., Neuhann, F., Glaser, N. et al. The path from a volunteer initiative to an established institution: evaluating 15 years of the development and contribution of the Lighthouse trust to the Malawian HIV response. BMC Health Serv Res 17, 548 (2017). 10.1186/s12913-017-2466-y

13. Ministry of Health and Population: Clinical Management of HIV in Children and Adults, National Guidelines, Malawi, 2022

14. https://www.omegadx.com/Portals/0/14266%20VISITECT%20CD4%20BROCHURE%20ISSUE%205_1.pdf, accessed 12.4.24

15. https://www.immy.com/package_inserts/cr2003/CR2003%20IFU%20(Int’l)%20-%20English.pdf, accessed 12.4.24

16. https://content.veeabb.com/1d09429b-8373-419f-8f1a-d28f9586863a/c5d7d9bb-803d-4456-b4ce-b4aeb385083b/c5d7d9bb-803d-4456-b4ce-b4aeb385083b_source_v.pdf

17. Fleiss, Joseph L., Jacob Cohen, and Brian S. Everitt. “Large sample standard errors of kappa and weighted kappa.” Psychological Bulletin 72.5 (1969): 323.

18. Nalintya E, Sekar P, Namakula OL, Tadeo KK, Kwizera R, Apeduno L, et al. The Diagnostic Performance of the Visitect Advanced Disease Point-Of-Care CD4 Platform: A Pragmatic, Mixed-Methods, Multisite Validation, Costing, and Qualitative Analysis. J Acquir Immune Defic Syndr. 2024 Dec 1;97(4):387–396. doi:10.1097/QAI.0000000000003505. PMID: 39159398; PMCID: PMC11732718.

19. Lechiile K et al. “Laboratory evaluation of the VISITECT advanced disease semiquantitative point-of-care CD4 test.” JAIDS Journal of Acquired Immune Deficiency Syndromes 91.5 (2022): 502–507.

20. Luchters S et al. “Field performance and diagnostic accuracy of a low-cost instrument-free point-of-care CD4 test (Visitect CD4) performed by different health worker cadres among pregnant women.” Journal of clinical microbiology 57.2 (2019): 10–1128.

21. Nakamaanya, O. Evaluation of the Diagnostic Validity of the HIV VISITECT CD4 Point-of-Care Rapid Diagnostic Test Using PIMA Analyzer as the Gold Standard in Uganda. Diss. Makerere University, 2023.

22. Ndlovu Z et al. “Diagnostic performance and usability of the VISITECT CD4 semi-quantitative test for advanced HIV disease screening.” Plos one 15.4 (2020): e0230453.

23. Otubu N, et al. “Comparison of Advanced HIV Disease identification using CD4 results from a semi-quantitative CD4 point of care test and CD4 flow cytometry in Nigeria.” JOURNAL OF THE INTERNATIONAL AIDS SOCIETY. Vol. 25, 2022.

24. Vidhyavathi V, et al. “Performance characteristics of an instrument-free point-of-care CD4 test (VISITECT® CD4) for use in resource-limited settings.” Journal of International Medical Research 48.9 (2020): 0300060520955028.

25. Gupta-Wright, Ankur, et al. “Rapid urine-based screening for tuberculosis in HIV-positive patients admitted to hospital in Africa (STAMP): a pragmatic, multicentre, parallel-group, double-blind, randomised controlled trial.” The Lancet 392.10144 (2018): 292–301.

26. Rick, Fernanda, et al. “Cryptococcal antigen screening by lay cadres using a rapid test at the point of care: a feasibility study in rural Lesotho.” PLoS One 12.9 (2017): e0183656.

27. MSF-Operational Center Brussels: Advanced HIV Disease: Every link in health care matters! Implementation of the ‘circle of care’ in rural Nsanje district, Malawi, December 2020. https://www.msf.org.za/sites/default/files/2023-02/Report_CapitalizationNsanjeMalawi_SAMU_2021_EN.pdf, accessed 29.5.2024

28. Heller, Tom, et al. “Implementing advanced HIV disease care for inpatients in a Referral Hospital in Malawi–demand, results and cost implications.” Annals of Global Health 88.1 (2022).

## References of publications included in the Forest plot

18. Kirungi R, Nabitaka V, Nuwagira A, Musoke A, Omar A, Moore A, Rathakrishnan D, Conroy J, Amole C, Namuwenge P, Nyegenye W, Okiira C, Kasone V: Implementation of the CD4 Advanced Disease Rapid Test: Lessons Learned from the Pilot Test in Uganda. Poster presented at 24th International AIDS Conference, AIDS 2022

